# Clinical Adherence to Concussion Practice Guidelines

**DOI:** 10.1101/2024.05.31.24308295

**Authors:** Ryan Pelo, Jonathan Godfrey, Leland E. Dibble

## Abstract

**Objective:** This study aimed to assess common barriers to adherence to the Physical Therapy Concussion/Mild Traumatic Brain Injury Clinical Practice Guidelines (PTCPG) and design an action plan to address these barriers.

**Setting:** Single University Health System

**Participants:** Electronic medical record (EMR) data were collected over an initial 6-month period and then a follow-up 6-month period after the interventions. The initial period yielded an average of 129 patients with a concussion diagnosis, and the follow-up period yielded an average of 331 patients.

**Design:** Through the knowledge-to-action framework, it was identified that providers were often unaware of current practice guidelines, and some had a low comfort level with the diagnosis. Subsequent action items to address these barriers included education sessions, modification of a concussion documentation template to better align with PTCPG recommendations, and the design of a pop-up within the electronic medical records that allowed providers to create a referral to a concussion treatment network.

**Main Measures:** Frequency calculations were performed based on whether the outcome measure was performed at any time during a concussion episode of care. Selection of outcome measures was determined by what outcome measures were available for use within the EMR and aligned with the PTCPG recommended system domains. Data was collected from the following providers: physical therapists, occupational therapists, sports medicine physicians and physical medicine & rehab physicians.

**Results:** From the initial EMR query to the follow-up, the following increases in use of outcome measures were seen with each outcome measure, Post-Concussion Symptom Scale (PCSS) = 10.0%, Cervical & Thoracic Screen = 6.0%, Vestibular Ocular Motor Screen (VOMS) = 12.6%, Modified Orthostatic Vital Signs (MOVS) = 18.9%, Buffalo Concussion Treadmill Test (BCTT) = 3.5%, Balance Error Scoring System (BESS) = -4.8%, Sensory Organization Test (SOT) = 0.9%.

**Conclusion:** The improvement in adherence to recommendations was attributed to a knowledge to action approach that combined education along with structural changes to electronic medical record documentation.

## INTRODUCTION

The 2020 Physical Therapy Concussion/Mild Traumatic Brain Injury Clinical Practice Guidelines PTCPG built upon previous clinical practice guidelines put forth in the 2016 Consensus Statement of Concussion in Sport (CSCS) and the 2018 Department of Veterans Affairs/Department of Defense Clinical Practice Guideline for the Management of Concussion-mild Traumatic Brain Injury (VA/DoD).^1–3^ The common thread across each of the guidelines is the proposal of mTBI sub-types designed to categorize impairments commonly experienced after a concussion. As guidelines have evolved, sub-types initially based on symptom presentation have transitioned to subtypes based on multi-modal body system pathophysiology.^4,5^ The recommendations within the Examination section of the PTCPG utilize the following body system domains: Cervical Musculoskeletal, Vestibulo-Oculomotor, Autonomic/Exertional and Motor Function, and recommends that following a concussion injury, each profile should be assessed if patient reported dizziness or headache is present. Given that dizziness and headache are reported in 80% and 91% of concussion injuries, respectively, all four domains should be assessed in most cases.^6,7^

Although practice guidelines are one means to disseminate information to providers about current best practice and provide a framework to guide clinical care, compliance rates across multiple health care disciplines and conditions have been found to only be about 50%.^8^ Given the specificity of the guidelines as to what a concussion examination should include, the goals of this study were to assess clinician performance over a 6-month period after the release of the PTCPG guidelines, to determine any inconsistencies in alignment of clinician performance with the PTCPG, and to determine and implement an action plan to bring clinical practice patterns closer to the PTCPG guidelines.

## METHODS

Data collection for this study was performed with approval from the University of Utah Institutional Review Board who deemed that this study did not qualify as Human Subject Research because it did not involve interaction or interventions with an individual or contain identifiable private information. The assessment of barriers and the determination of an action plan was guided by the knowledge to action framework which guides knowledge translation through a series of phases that continually adjust and adapt in order to address barriers to learning.^9^

### Identify Problem/Identify, Review, Select Knowledge

Study data was collected through the University of Utah Health System electronic medical records request. The date range of the initial data pull was April 1^st^, 2020 – September 30^th^, 2020. Due to this being during the COVID pandemic and the use of telehealth appointments being utilized across the healthcare system, only in-person visits were included in this study as most of the assessments needed to be performed in-person. Providers included in the data request were meant to replicate the multi-disciplinary approach to concussion care within the University of Utah Health System and contained: physical therapists, occupational therapists, and physicians within either the Sports Medicine or Physical Medicine & Rehabilitation Departments. The break down by percentage of providers in each discipline were: 67% physical therapists, 8% occupational therapists, 20% sports medicine physicians, and 5% physical medicine & rehab physicians. Concussion diagnosis was determined by the following ICD-10 codes: S06.0* - Concussion, F07.81 - Post-Concussive Syndrome, S06.2X9D - Mild Traumatic Brain Injury with Loss of Consciousness unspecified, S06.0X0A - Mild Traumatic Brain Injury with Loss of Consciousness, S06.0X1A - Mild Traumatic Brain Injury with Loss of Consciousness less than 30min, and G44.309 - Post-Traumatic Headache.

Selection of outcome measures was determined by what outcome measures were available for use within the EMR and aligned with the PTCPG recommended system domains. Outcome measures that met this criteria included: Post-Concussion Symptom Scale (PCSS)^10^, a Cervical & Thoracic Screen, Vestibular Ocular Motor Screen (VOMS)^11^, Modified Orthostatic Vital Signs (MOVS)^12^, Buffalo Concussion Treadmill Test (BCTT)^13^, Balance Error Scoring System (BESS)^14^, and Sensory Organization Test (SOT)^15^.

Frequency calculations were performed based on whether the outcome measure was performed at any time throughout a concussion episode of care. A concussion episode of care was deemed to include all visits from physical therapists, occupational therapists, sports medicine physicians and physical medicine & rehab physicians related to a single concussion incident. This was done to attempt to reduce the scenario in which a physical therapist may have not performed an outcome measure because it had already been performed by a physician or occupational therapist. If a patient were to have multiple concussions during this period, each concussion incident was considered a new episode of care and outcome measures performed during a previous concussion episode of care did not count toward performance during the new concussion (Table 1).

**Table 1:** Categorization of outcome measures based on 2020 Physical Therapy Concussion/Mild Traumatic Brain Injury Clinical Practice Guidelines (PTCPG) body system domains.^3^.

| PTCPG Domains |  | Assessments |
| --- | --- | --- |
| Cervical Musculoskeletal | examine the cervical and thoracic spines for potential sources of musculoskeletal dysfunction | Cervical & Thoracic Screen |
| Vestibulo-oculomotor | examine vestibular and oculomotor function<br>examine for benign paroxysmal positional vertigo (BPPV), if indicated | VOMS |
| Autonomic/Exertional | evaluate heart rate and blood pressure in supine, sitting, and standing positions.<br>perform symptom-guided, graded exertional tolerance test | MOVS, BCTT |
| Motor Function | examine motor function impairments, including static balance, dynamic balance, motor coordination and control, and dual/multitasking | BESS, SOT |
| Description of testing recommendations within each domain and categorization of measured that were assessed into each domain.<br>**Abbreviations:** BPPV = Benign paroxysmal positional vertigo, PCSS = Post-Concussion Symptom Scale, VOMS = Vestibular Ocular Motor Screen, BESS = Balance Error Scoring System, SOT = Sensory Organization Test, MOVS = Modified Orthostatic Vital Signs, BCTT = Buffalo Concussion Treadmill Test. |  |  |

### Adapt Knowledge to Local Context/Assess Barriers to Knowledge Use/Select, Tailor, Implement Interventions

To identify and address the contributing barriers to reduced guideline adherence and to create action items to improve best practice, a 20-member Concussion Rehabilitation Committee consisting of physical therapists, occupational therapists, sports medicine physicians and physical medicine & rehab physicians across the university-based health system was created. Discussion within the committee was then taken back to individual discipline groups within the Sports Medicine and Physical Medicine and Rehabilitation Departments. Through discussion and informal survey, it was determined that two primary issues consistently emerged as reasons why clinicians did not utilize outcome measures as advocated in the guideline recommendations: 1) Clinicians were unaware or unable to keep up with current concussion best practice guidelines, 2) Not all clinicians seeing individuals post-concussion had an appropriate comfort level with the diagnosis. Provided with this information, the committee established a 2-step approach to address the barriers to guideline adherence which included an education component along with an adaptation of the electronic medical record system to help facilitate long-term compliance (Figure 1).

**Figure 1.**
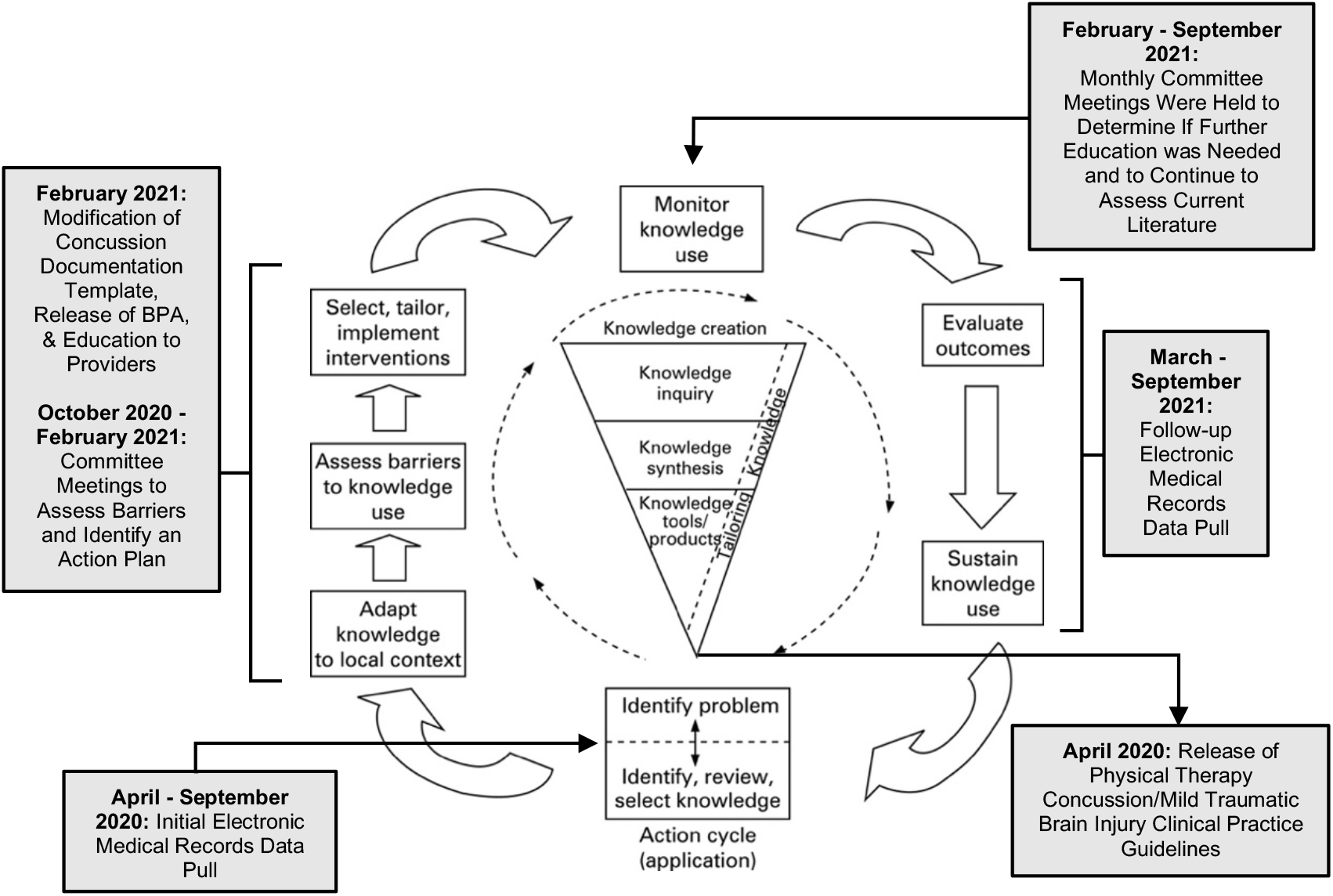
Knowledge-to-action framework diagram with timeline of current study stages of framework. Image and framework design adapted from Graham et al., 2006. **Abbreviations:** BPA = Best Practice Alert

To address issue 1), a concussion documentation template within the electronic medical records was modified to be more user friendly and to create a workflow in line with the PTCPG guideline recommendations. Clinical education was provided through educational inservice meetings and grand rounds sessions to both the committee and to provider groups throughout the health system. Using this educational process as a foundation, a concussion treatment network was created. This network included clinicians that represented all outpatient rehabilitation sites in the Health System.

To address issue 2), a process was created within the electronic medical record system that created a Best Practice Alert (BPA) pop-up when a concussion diagnosis was entered within a patient’s medical record. The BPA would allow for the ability to guide a referral to a concussion treatment network provider. This referral option allowed providers that were not comfortable treating concussion a pathway to provide patients with the appropriate care through a referral to providers trained in concussion care.

### Monitor Knowledge Use

Monthly committee meetings were held to determine the effectiveness of the educational interventions through observations of practice patterns and whether further inservice meetings, grand rounds or other education opportunities were necessary. The committee also continued to review current literature to assess best practice for concussion and decided if any updates were needed.

### Evaluate Outcomes/Sustain Knowledge Use

To determine the effect of our 2-step process to improve clinician adherence to concussion guideline recommendations, a follow-up electronic medical record data pull was performed (date range: March 1^st^, 2021 – August 31^st^, 2021). Overall numbers of referrals were also gathered. Results were analyzed and compared against the initial data pull results.

An additional sub-committee was created that was tasked with regularly reviewing current research to that informed recommended concussion practices. This review process was intended to provide the opportunity to iteratively update the electronic medical record concussion documentation template and to continue to disseminate current information to clinicians.

## RESULTS

The same departments within the University of Utah Health System, which contained: physical therapists, occupational therapists, and physicians within either the Sports Medicine or Physical Medicine & Rehabilitation Departments, were assessed as in the initial EMR query. A substantial increase in patient referrals to concussion treatment network providers were observed. The initial records request returned an average of 129 patient visits per month and the follow-up records request returned an average of 331 patient visits per month. Usage of the concussion documentation template also increased from 44 providers initially to 67 providers after the educational interventions and documentation changes were implemented.

The initial EMR query found the following performance in each domain, Post-Concussion Symptom Scale (PCSS) = 44.0%, Cervical & Thoracic Screen = 17.8%, Vestibular Ocular Motor Screen (VOMS) = 39.5%, Modified Orthostatic Vital Signs (MOVS) = 13.4%, Buffalo Concussion Treadmill Test (BCTT) = 10.2%, Balance Error Scoring System (BESS) = 14.0%, Sensory Organization Test (SOT) = 15.9% (Table 2).

**Table 2:** Frequency calculations of assessments performed during a patient’s course of treatment for concussion.

|  |  |  |  |  |  |
| --- | --- | --- | --- | --- | --- |
| Profiles | % of Patients Pre | % of Patients Post | Assessments | % of Patients Pre | % of Patients Post |
| Symptoms | 44.0 | 54.0 | PCSS | 44.0 | 54.0 |
| Vestibulo-oculomotor | 39.5 | 52.1 | VOMS | 39.5 | 52.1 |
| Motor Function | 29.9 | 26.0 | BESS | 14.0 | 9.2 |
|  |  |  | SOT | 15.9 | 16.8 |
| Cervical Musculoskeletal | 17.8 | 23.8 | Cervical & Thoracic Screen | 17.8 | 23.8 |
| Autonomic/Exertional | 23.6 | 46.0 | MOVS | 13.4 | 32.3 |
|  |  |  | BCTT | 10.2 | 13.7 |
| % of patients indicates the number of patients that had documentation of their domain and assessments listed in their medical record. |  |  |  |  |  |
| **Abbreviations:** PCSS - Post-Concussion Symptom Scale, VOMS - Vestibular Ocular Motor Screen, BESS - Balance Error Scoring System, SOT - Sensory Organization Test, MOVS - Modified Orthostatic Vital Signs, BCTT - Buffalo Concussion Treadmill Test. |  |  |  |  |  |

After the educational interventions and documentation changes were implemented, the Motor Function (BESS + SOT) profile was the only profile that showed a decrease in performance. From the initial EMR query to the follow-up, providers demonstrated the following changes in use for each outcome measure, Post-Concussion Symptom Scale (PCSS) = +10.0%, Cervical & Thoracic Screen = +6.0%, Vestibular Ocular Motor Screen (VOMS) = +12.6%, Modified Orthostatic Vital Signs (MOVS) = +18.9%, Buffalo Concussion Treadmill Test (BCTT) = +3.5%, Balance Error Scoring System (BESS) = -4.8%, Sensory Organization Test (SOT) = +0.9% (Table 2).

## DISCUSSION

With the objective of improving the care process for individuals participating in post-concussion rehabilitation, our health system examined provider performance prior to and following a knowledge translation effort. The process of analyzing health system electronic medical records provided us with knowledge about current clinical practice and how it aligned with practice guidelines. The information gained guided the process of educating providers, creating a concussion rehabilitation referral network, modifying the concussion documentation template, and the creation of a best practice alert pop-up. The creation of a best practice alert was felt to be unique in that it did not simply rely on education of providers but put in place changes to the electronic medical record system that would direct providers toward appropriate referral options.

Studies examining symptom subtypes report that dizziness and headache are present in 80% and 91% of concussion injuries, respectively.^6,7^ Assuming the concussions referred for rehabilitation care in our system possess similar symptomology to what is reported in the literature, it would be expected that these examination tools relevant to the concussion subtype would have been performed more frequently than the less than 25% of concussion cases that was observed prior to our knowledge translation efforts. This discrepancy is what guided the Concussion Rehabilitation Committee through discussions and identified the lack of clinician awareness and the lack of clinician comfort as the barriers to greater guideline adherence. Interventions developed using a knowledge to action framework contributed to substantial increases in the use of examination tools consistent with clinical practice guideline adherence and increases in concussion rehabilitation referrals.

While there was a general increase in outcome measure utilization, it is unclear why there was a drop in the use of outcome measures within the Motor Function system domain. One potential cause is the static nature of the postural control measures in our template. Both the BESS and the SOT only ask the subject to stand still and control center of mass motion. Given the wide variety of outcome measures that assess dynamic stability (such as the FGA and HiMAT), there is a need to expand the options that are provided within the concussion documentation template, as there is the potential that providers were using alternate outcome measures to the BESS and the SOT that were not captured in this analysis. As part of continued evaluation of the care process, an increase in the spectrum of motor function measures that are included in the documentation template is on-going.

### Limitations and Directions for Future Modifications to the Care Process

Although we showed improved provider adherence with clinical practice guidelines following our knowledge translation efforts, these results have several limitations. The outcome measures we assessed were limited to what was already included within the concussion documentation template within the EMR. This restricted our frequency calculations, and it is quite possible that additional outcomes measures could have been performed outside of the documentation template and therefore were not included in our analysis.

We also recognize that there could be some redundancy in domains that have multiple measures, as during a single episode both measures within the domains could have been performed. This would have counted as two performances in the total numbers and therefore would have driven up the percentage of performance within the total sample of concussion episodes. Although the general understanding of frequency of performance of outcome measures within a domain would not have changed, this correction is something to consider with future data collection.

The consistency of clinician documentation within the electronic medical record’s concussion documentation template could have also influenced our performance frequency numbers. Consistent documentation within the specific concussion documentation template has been deemed to be an action item going forward.

Given the time period of data collection coinciding with the COVID pandemic, it is likely that visit numbers were impacted and overall care throughout the health care system was adjusted in many ways. To account for this, only in-person visits were included in this study as most of the assessments needed to be performed in-person.

## CONCLUSION

Through an approach that utilized a multidisciplinary Concussion Rehabilitation Committee coupled with provider education and EMR modifications, we influenced the care process and improved provider adherence to concussion rehabilitation clinical practice guidelines. While this process improved the scope of examination of concussion sub-types, it did not examine treatment. Future efforts should explore treatment processes.

## Data Availability

All data produced in the present study are available upon reasonable request to the authors

## ACKNOWLEDGEMENTS

The authors would like to thank the University of Utah Concussion Rehabilitation Committee for their work in trying to continually improve patient care.

